# The global burden of yellow fever

**DOI:** 10.1101/2020.10.14.20212472

**Authors:** Katy A. M. Gaythorpe, Arran T. P. Hamlet, Kevin Jean, Daniel Garkauskas Ramos, Laurence Cibrelus, Tini Garske, Neil M. Ferguson

**Affiliations:** WHO Collaborating Centre for Infectious Disease Modelling, MRC Centre for Global Infectious Disease Analysis, Abdul Latif Jameel Institute for Disease and Emergency Analytics (J-IDEA), Imperial College London, Praed Street, London, UK; Maître de conférences,Laboratoire MESuRS - Cnam Paris, 292 rue Saint Martin, 75003 Paris; Secretariat for Health Surveillance, Brazilian Ministry of Health, Brasilia, Brazil; World Health Organisation, Geneva, Switzerland

## Abstract

**Background:** Yellow fever (YF) is a viral haemorrhagic fever endemic in tropical regions of Africa and South America. Current intervention policies, namely the Eliminate Yellow fever Epidemics (EYE) strategy are actioned through vaccination. However, the stockpiles and production mean that vaccination can be in short supply. As such, intervention strategies need to be optimised; one of the tools for doing this is mathematical modelling.

**Methods:** We fit a generalised linear model of YF reports to occurrence data available from 1987 to 2019 in Africa and South America and available serology survey data to estimate the force of infection across the continents. Then, using demographic and vaccination data, we examine the impact of interventions.

**Findings:** We estimate that in 2018 there were approximately 51,000 (95%CrI [31,000 - 82,000]) deaths due to YF in Africa and South America. When we examine the impact of mass vaccination campaigns in Africa, these amount to approximately 10,000 (95%CrI [6,000 - 17,000]) deaths averted in 2018 due to mass vaccination activities in Africa; this corresponds to a 47% reduction (95%CrI [10% - 77%]).

**Interpretation:** We find that the majority, 92% (95%CrI [89% - 95%]), of global burden occurs in Africa and that mass vaccination activities have significantly reduced the current deaths per year due to YF. This methodology allows us to evaluate the effectiveness of vaccination campaigns past, present and future and illustrates the need for continued vigilance and surveillance of YF.

**Funding:** BMGF and MRC

## Introduction

Yellow fever (YF) is a flavivirus endemic in tropical regions of Africa and South America. In Africa, it is the third most commonly reported type of disease outbreak^1^ in the Americas, YF produces extensive epizootics in NHP and outbreaks of human cases. It is vaccine preventable, with a safe and effective vaccine available since the 1930s that has been introduced into the Expanded Programme on Immunisation (EPI) of many countries^2^. YF is transmitted by numerous vectors including *Aedes* spp. and *Haemogogus* spp. in Africa and the Americas respectively. A component of the sylvatic reservoir system is in non-human primates (NHPs) and as a result of this, YF cannot be eradicated. The clinical course of YF infection leads to a variety of non-specific symptoms with severe infections potentially exhibiting fever, nausea, vomiting, jaundice and hemorrhaging which can result in death^3^.

The transmission dynamics of YF have numerous components. There are three main “cycles”, urban, and sylvatic and intermediate. The sylvatic cycle is said to be the driver of most reported transmission with infection occurring mainly between NHPs mediated by tree hole breeding mosquitos. These vectors are diurnal and feed mostly on NHP; however, people can be infected if they encroach on this cycle through occupational or recreational activities^3^. In South America, this accounts for the majority of cases with potentially large outbreaks; a recent YF season saw over 1,000 cases in Brazil alone^4^. The urban cycle of YF is less common but the outbreaks have the potential to be devastating. Whilst urban outbreaks have largely been eradicated in South America^5^, they still occur in Africa with a recent urban outbreak, in Angola and the Democratic Republic of the Congo, causing 962 reported cases, though this is thought to be a fraction of the actual transmission^6^. Large outbreaks as a result of urban transmission are due to the combination of densely populated urban areas, and large populations of *Ae. Aegypti*, which bites humans preferentially and breeds rapidly in urban environments^7^. The World Health Organisation (WHO) developed the Eliminate Yellow Fever Epidemics (EYE) strategy in order to eliminate urban YF outbreaks by 2026^8^. The intermediate cycle currently only occurs in Africa when tree-hole breeding anthropophilic *Aedes* reach particularly high densities^3^.

Control of YF is primarily through vaccination with no specific anti-viral treatment available. The, 17D vaccine is live attenuated and was developed in 1936^9^. The vaccine is considered safe; estimates of adverse events have ranged up to 0.6/100,000 doses, with reactions are generally mild^10^. Immunity due to yellow fever vaccination is suggested to be lifelong with recommendations recently updated to reflect this^11^. Efficacy is also thought to be high with recent estimates suggesting serological response was 97.9% (95%CrI [82.9 - 99.7])^12^. An issue with the vaccine is production and corresponding stockpiles. As the vaccine is live, production is slow which has led to vaccine shortages for large outbreaks and for travellers^13^. As a result of this, fractional dosing has become a recommendation in outbreak settings^14^.

Due to limited vaccine supply, efficient planning of interventions is vital to avoid large outbreaks. To facilitate this, robust estimates of disease burden and projections of future dynamics are key. Previous studies have focused on evaluating historical vaccine impact and projecting future potential impact. Garske et al. produced vaccine impact estimates for the African endemic region focusing on mass vaccination campaigns until 2013^15^. They found that mass vaccination activities have averted burden by 57% (95%CI [54 - 59]) in countries where they took place, accounting for 27% (95%CI [22 - 31]) of the burden across the region. More recently, Shearer et al. examined the impact of vaccination globally, across Africa and South America, and found that (all) vaccination activities averted between approximately 94,0000 and 119,000 cases each year^16^. In this study, we refine and extend the model of Garske et al. to encompass new geographic regions: South America; new data: on occurrence, serology, vaccination and NHPs, and produce updated estimates of burden and vaccine impact for YF. In the following sections we describe the new data and modelling approach, particularly focusing on the updated model of yellow fever occurrence. Then we present results of our projected transmission intensity considering uncertainty from estimation and the structural uncertainty of the models. Finally, we present burden estimates and reassess the impact of mass vaccination activities in Africa.

## Methods

We expand an existing framework for Africa to account for transmission in South America. Countries are included if they have been listed as at-risk, endemic or potentially at risk for YF^8^. We fit a generalised linear model of YF reports to occurrence data available from 1984 to 2019, see figure 1. These occurrences denote whether YF has been reported at all in that time, irrespective of number of cases. This provides a probability of YF report. The number of infections required to produce a probability of report is then estimated using available serology survey data which provides, in specific locations, an independent estimate of the force of infection. These individual estimates allow us to calculate a probability of detection over the observation period which we may then use to provide force of infection estimates for the entire region. Finally, using demographic and vaccination data, we can calculate the burden in all provinces.

**Figure 1:**
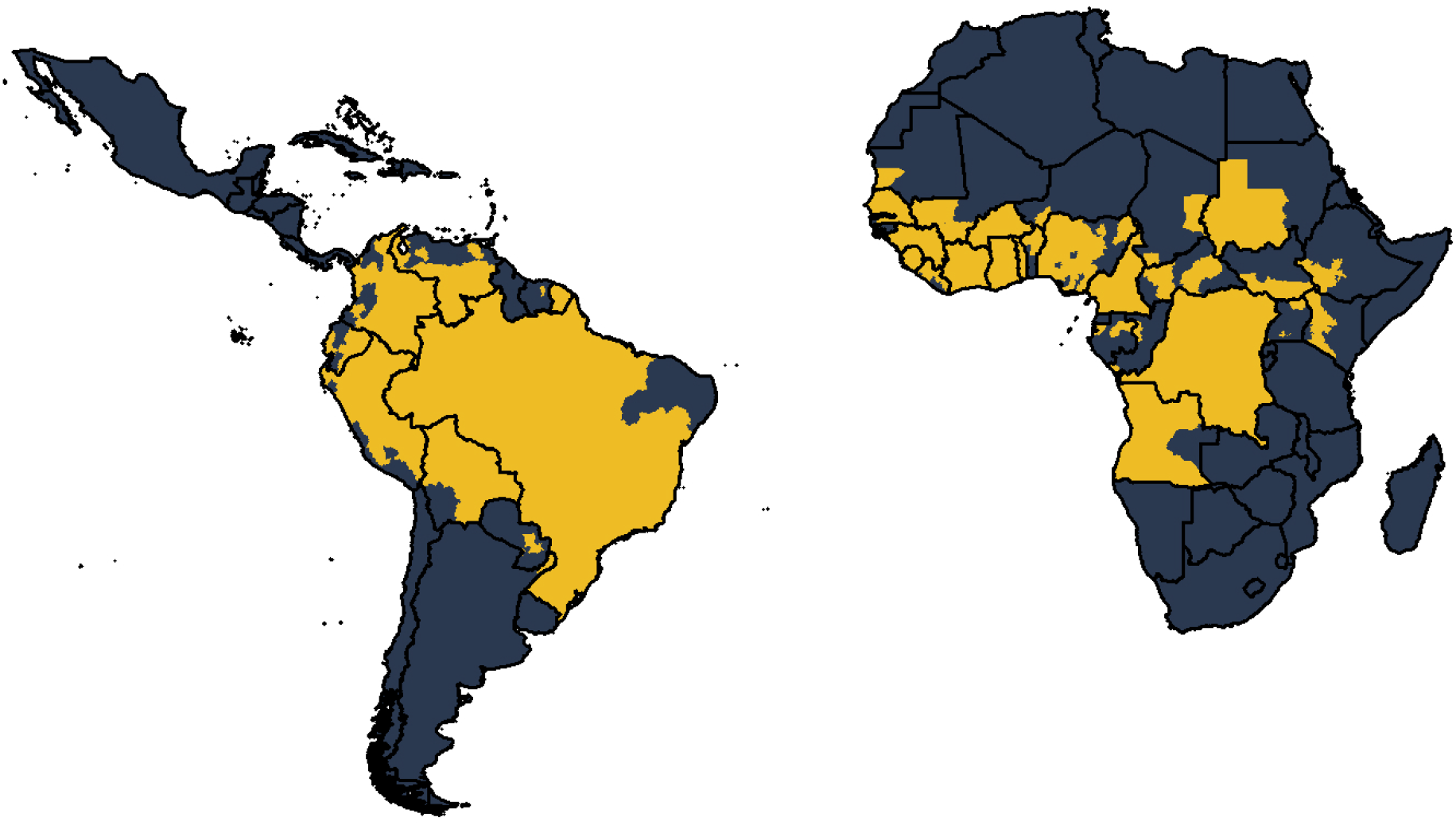
Global occurrence of yellow fever at admin level 1. Occurrence since 1984 is shown in yellow.

All data and models are described in the supplementary material.

All analyses, estimation and the original draft of the manuscript were performed in R version 3.5.1.

All data was from secondary sources and ethics approval was thereby not required.

## Results

### Regression model fitting and variable selection

All of the models included log (surveillance quality) and country factors for which country specific surveillance information was not available. A total of 50 covariates were considered, all of which were significantly related to the data with threshold p = 0.1. These were clustered into 39 groups leading to approximately 2.03 × 10^46^ model permutations. Following the use of a step function based on BIC, we reduce this number to 13 covariates and retain the 20 best models including these, shown in tables S1 and S2.

Similar to Garske et al., all of the 20 best fitting models included the log of population size, relating the probability of a report with the human population. All 20 models also included the temperature suitability at mean temperature which will limit the models in areas where the temperatures are too extreme to sustain vector transmission dynamics. The species richness of three NHP families were included in all variants. These were aggregated to form a variable `aggregate_family` in order to balance the contribution of the NHP host with the competence of vectors and human dynamics and populations. All model covariates are shown in figure 2. Covariates such as *Ae. Aegypti* occurrence, temperature range, altitude and barren, cropland, shrubland and water body land cover were only included in some of the best models.

**Figure 2:**
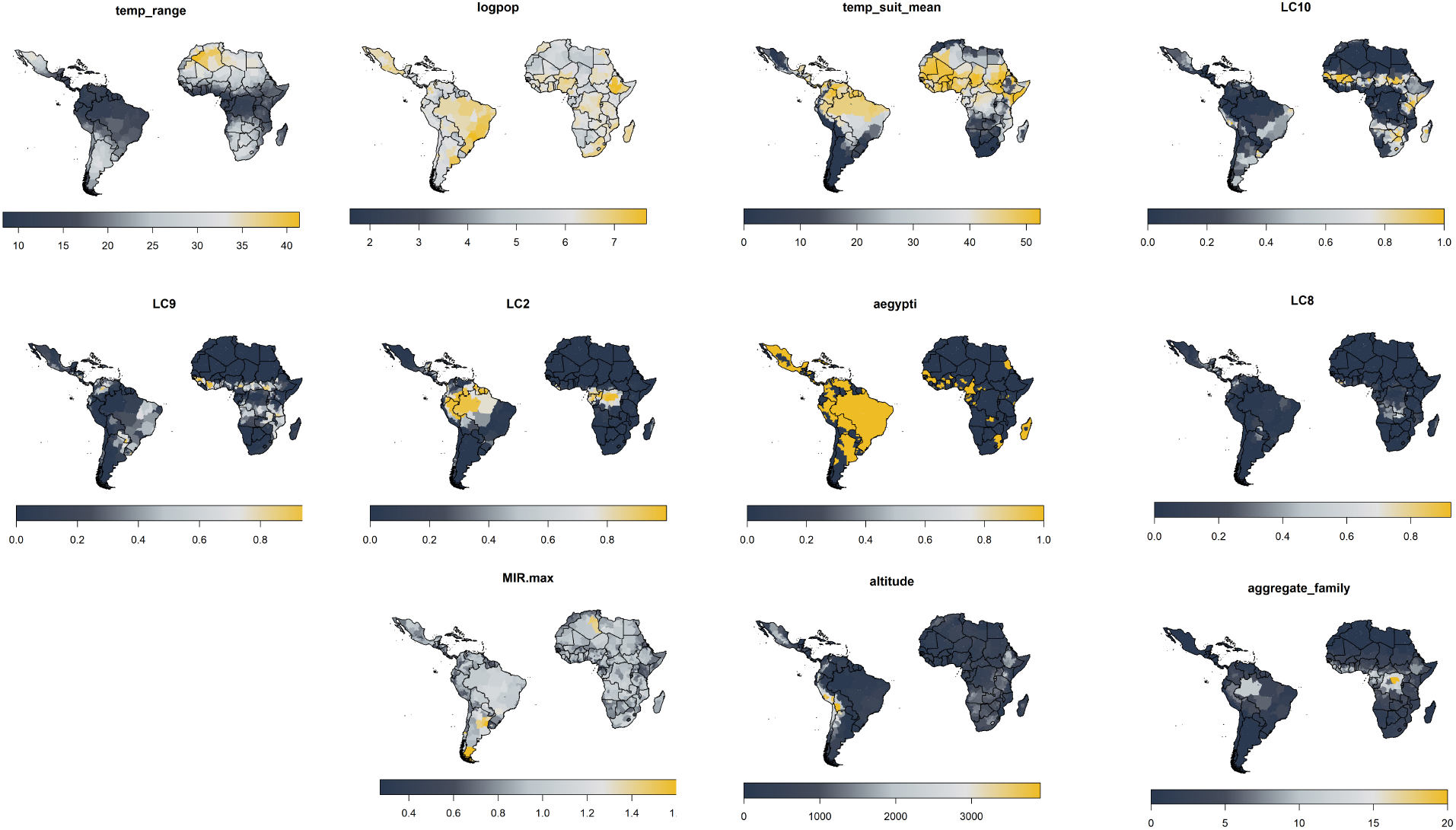
Maximal list of model covariates. Aggregate family is the sum of all NHP species richness in table 1 and will vary as families are included/ excluded.

The 20 best fitting models were also fit with full MCMC and the AUC calculated. These are shown in figure 3. The AUC of model variants are very similar with variant 6 generally the best. All model variants exceed the AUC of 17 which had point estimate of 0.916.

**Table 1:** Potential deaths and severe infections per year in Africa and South America from ensemble model projections.

| Continent | Year | Severe<br>infections,<br>median | Severe<br>infections,<br>95%CrI<br>low | Severe<br>infections,<br>95%CrI<br>high | Deaths,<br>median | Deaths,<br>95%CrI<br>low | Deaths,<br>95%CrI<br>high |
| --- | --- | --- | --- | --- | --- | --- | --- |
| Africa | 1995 | 102,972 | 62,162 | 160,700 | 48,474 | 28,672 | 76,998 |
| Africa | 2005 | 122,101 | 74,915 | 192,773 | 57,182 | 34,446 | 90,736 |
| Africa | 2013 | 98,148 | 62,083 | 150,953 | 45,973 | 28,680 | 72,380 |
| Africa | 2018 | 100,952 | 63,001 | 158,362 | 47,318 | 29,162 | 74,981 |
| Americas | 1995 | 14,349 | 6,528 | 26,016 | 6,652 | 3,026 | 12,577 |
| Americas | 2005 | 10,254 | 4,988 | 18,436 | 4,827 | 2,265 | 8,779 |
| Americas | 2013 | 8,559 | 4,264 | 15,043 | 3,999 | 1,969 | 7,162 |
| Americas | 2018 | 8,331 | 4,306 | 14,608 | 3,883 | 1,971 | 7,033 |

**Figure 3:**
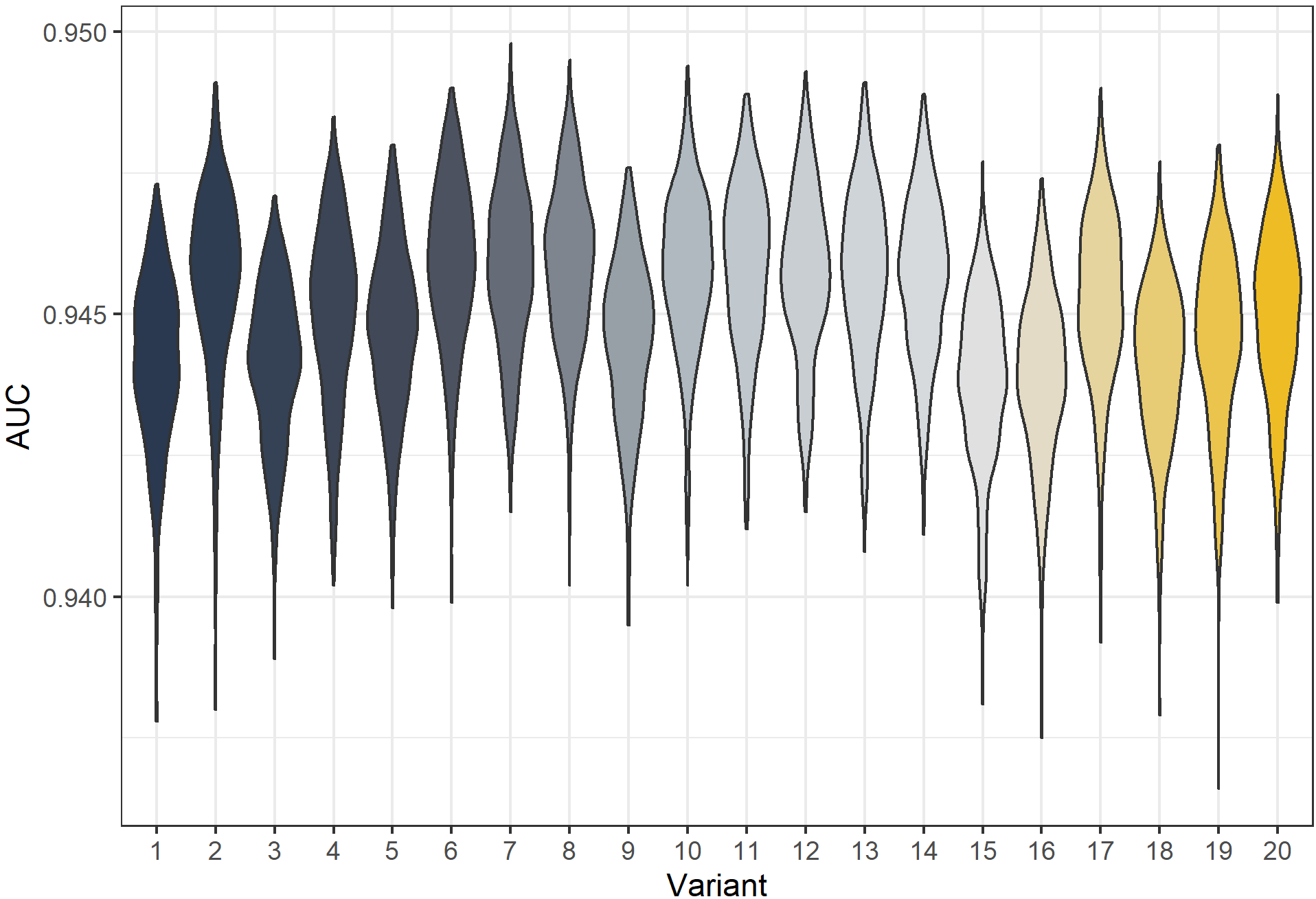
Posterior predicted area under the curve (AUC) for all model variants. The AUC are calculated for 500 samples from the posterior of each model variant.

### Yellow fever occurrence

We predict YF occurrence over the observation period in figure 4. These ensemble predictions indicate high probabilities of report in the Amazon region of Brazil and West Africa. Note the results shown also include a measure of surveillance effort emphasizing countries such as Angola.

**Figure 4:**
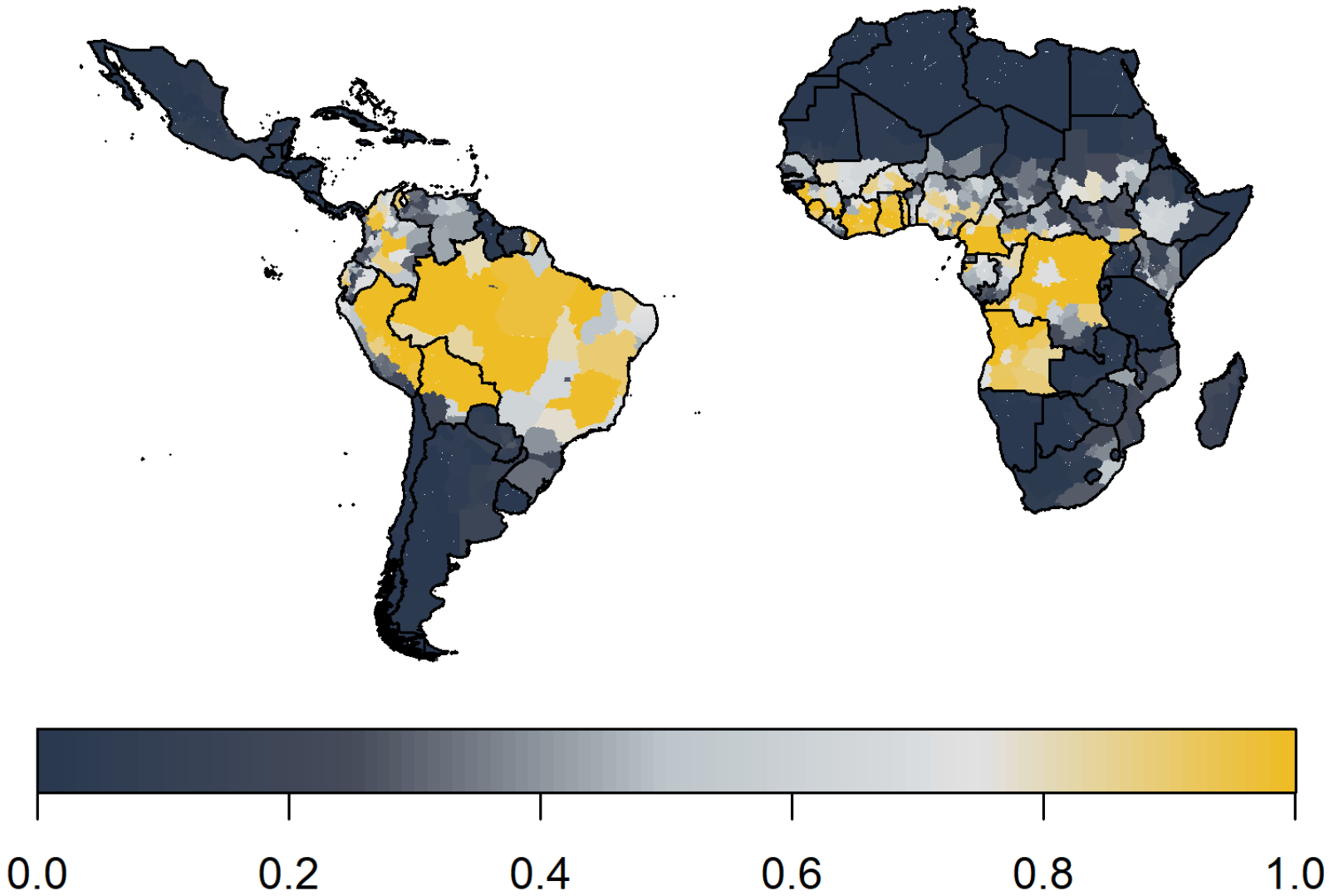
Median posterior predicted probability of a YF report from ensemble predictions of the 20 best GLMs.

### Seroprevalence

The model prediction captured the wide range of transmission intensity, see figure 5. However in certain conditions such as in Kenya area 1, the fit is affected by uncertainty concerning the vaccination status of included individuals. These results show similar qualitative fits to 17. Indeed there are only two additional included studies, those found in Kenya of 18.

**Figure 5:**
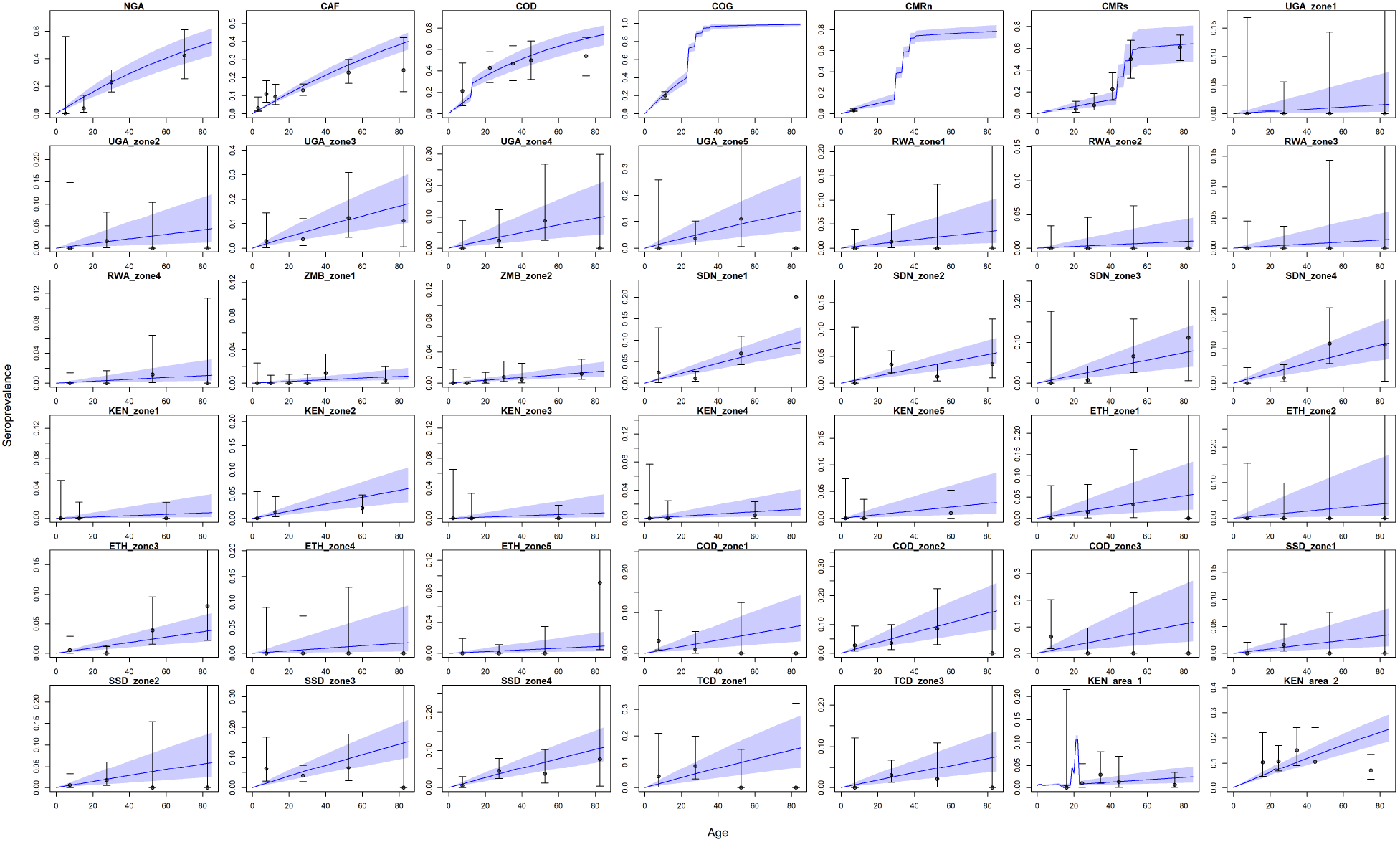
Seroprevalence predictions for each serological survey. Central blue line indicates median posterior predicted seroprevalence; blue area indicates 95%CrI.

### Transmission intensity

In figure 6 we show the median ensemble predictions of transmission intensity. In comparison to Garske et al. the force of infection in West Africa is slightly lower and provinces in the Democratic Republic of the Congo (DRC) are highlighted as areas of high transmission intensity. However, the main area highlighted is that of Amazon in Brazil. See supplementary material for the coefficient of variation between 100 samples from each of the 20 models, sampled equally; this further highlights areas of low transmission intensity such as the Sahara having higher degrees of uncertainty.

**Figure 6:**
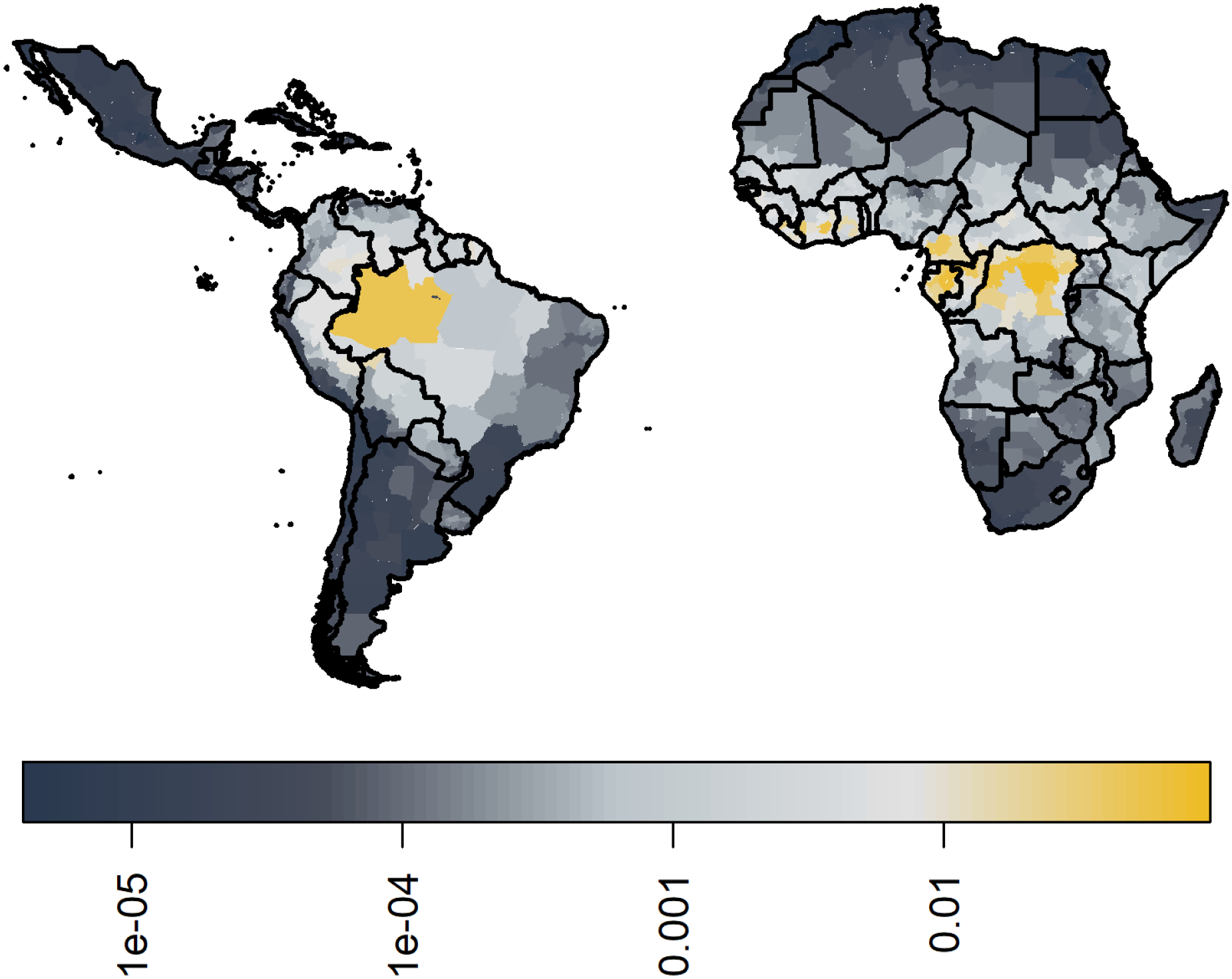
Median posterior predicted force of infection from ensemble predictions of the 20 best GLMs.

### Burden

The annual potential number of deaths and severe infections are shown in table 1 with deaths per country in 2018 given in figure 7. We estimate that in 2018 there were approximately 109,000 (95%CrI [67,000 - 173,000]) severe infections and 51,000 (95%CrI [31,000 - 82,000]) deaths due to YF in these two regions. Burden is distributed unevenly between countries and continents. The highest burden is seen in the DRC due to a high force of infection and low vaccination coverage. In contrast Brazil sees the fourth highest burden purely due to high force of infection in the Amazon region rather than low vaccination coverage. The majority of the burden occurs in Africa which holds for all years shown.

**Figure 7:**
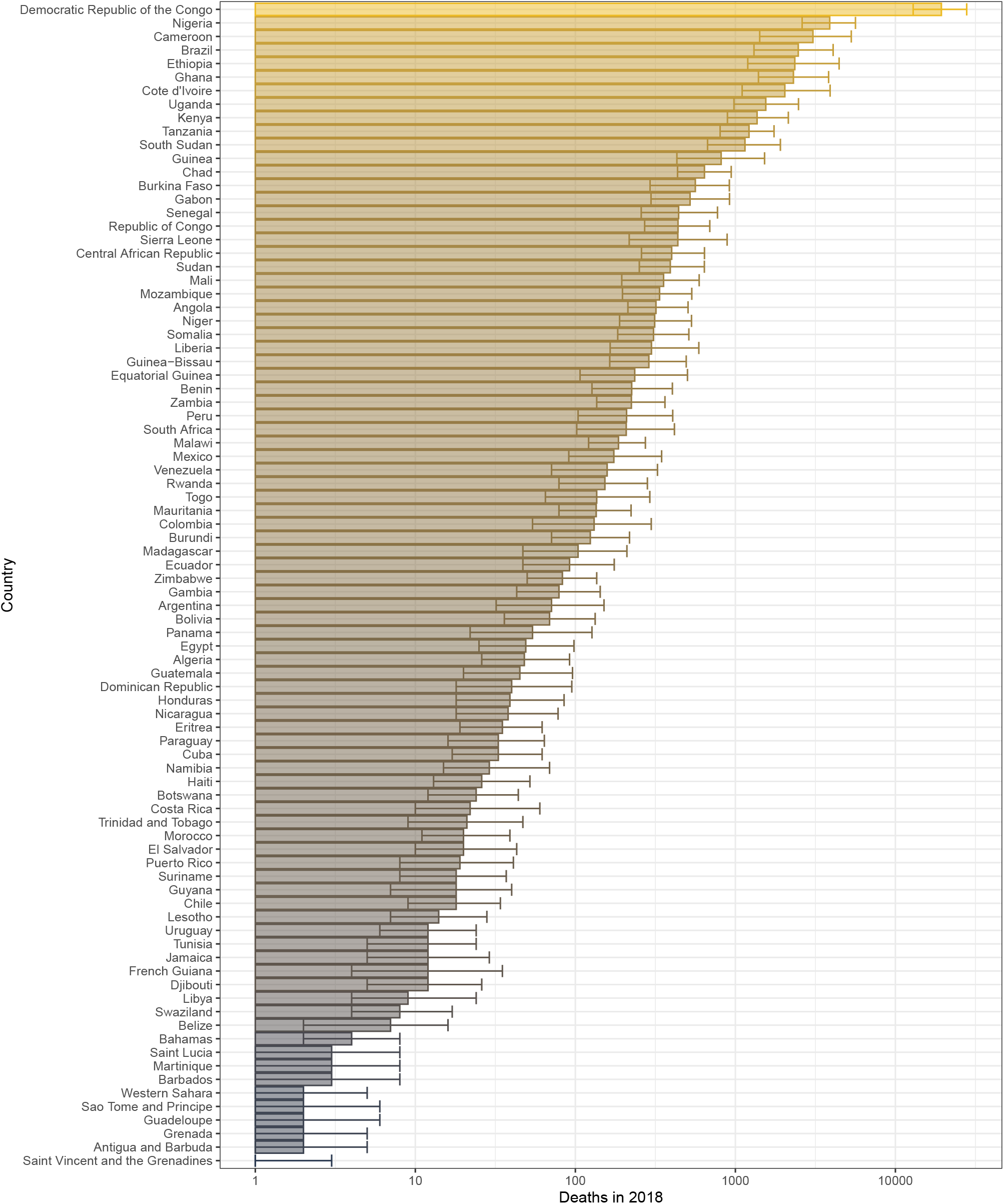
Posterior predicted potential deaths per country in 2018 from the ensemble model projections.

### Impact of mass vaccination campaigns

It has been shown that mass vaccination campaigns can produce long lasting effects on disease burden. We examine the effects of mass vaccination activities from 2006 until 2019 in countries in Africa, similar to 15, in figure 8 and in the supplementary material for 2013. We find large reductions in all countries with mass vaccination campaigns. In 2018, the largest reductions are of approximately 73% (95%CrI [64 - 79]) in Benin, 73% (95%CrI [60 - 81]) in Togo and 61% (95%CrI [52 - 68]) in Liberia, see supplementary material for further values.

**Figure 8:**
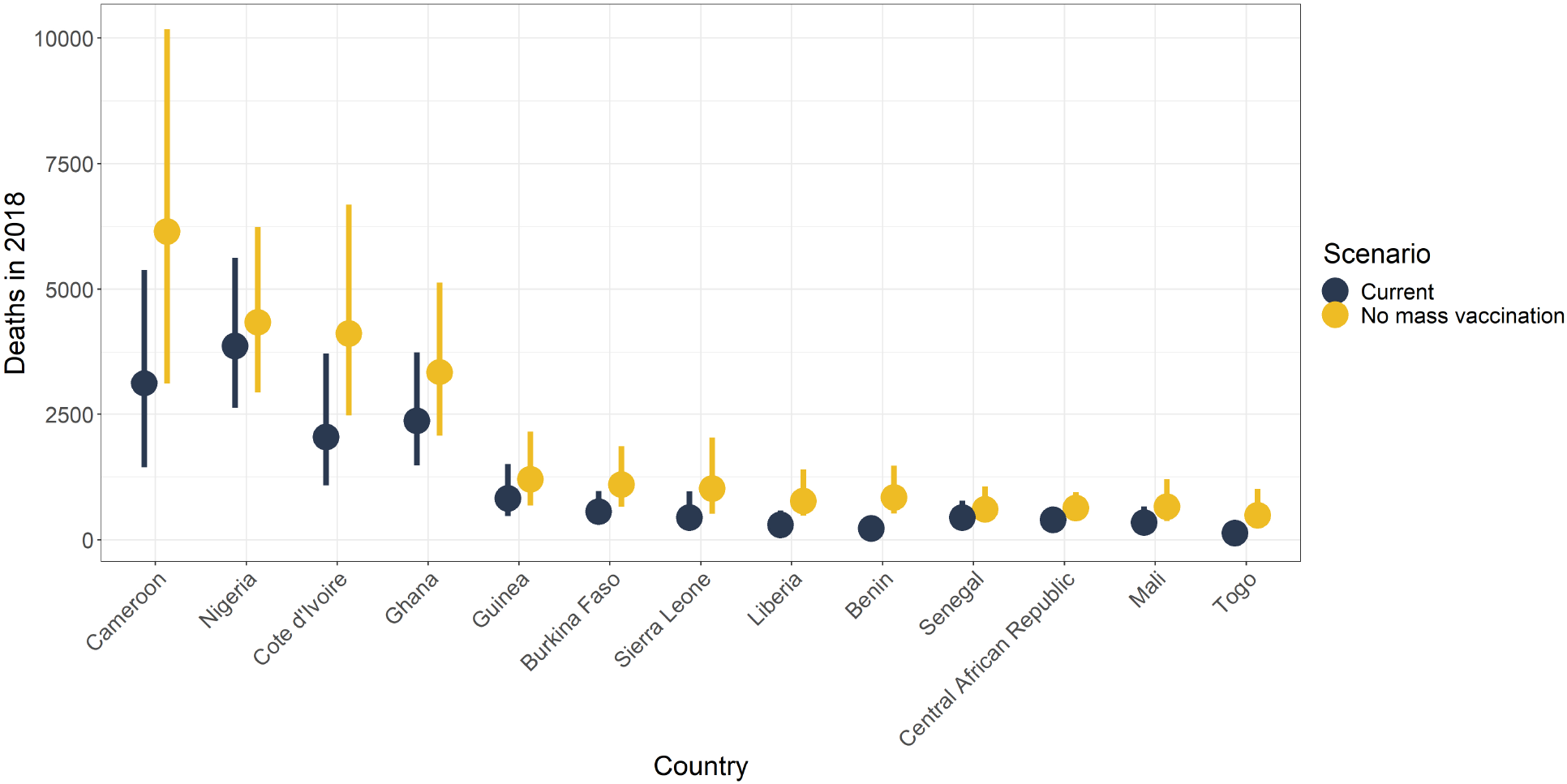
Median posterior predicted deaths averted for 2018 by country. Yellow represents the number of deaths without mass vaccination campaigns since 2006 and black represents deaths with current vaccination coverage levels. The points denotes median and the line shows the 95% credible interval.

Overall, the reductions in the number of deaths per year are substantial, shown in table 2. These amount to approximately 10,000 (95%CrI [6,000 - 17,000]) deaths averted in 2018 due to mass vaccination activities in Africa; corresponding to a 47% reduction (95%CrI [10 - 77]) in deaths.

**Table 2:** Deaths averted per year due to mass vaccination activites occurring from 2006 onwards in Africa.

| Year | Median deaths averted | Deaths averted, 95%CrI<br>low | Deaths averted, 95%CrI<br>high |
| --- | --- | --- | --- |
| 2013 | 11,414 | 6,400 | 19,369 |
| 2018 | 10,140 | 5,781 | 17,307 |

## Discussion

In this study we deveoped models yellow fever transmission in Africa and South America. We calculated disease burden in terms of severe infections (or cases) and deaths from an ensemble of best-fitting generalised linear models (GLMs) of yellow fever reports between 1987 and 2018 coupled to mechanistic models of seroprevalence. We used this approach to evaluate the impact of mass vaccination campaigns in Africa as well as produce updated burden estimates of yellow fever in endemic regions.

We estimate that there are between 63,000 - 158,000 severe infections of YF in Africa, resulting in 29,000 - 75,000 deaths. In South America, we estimate there are 4,000 −15,000 severe infections, resulting in 2,000 - 7,000 deaths. These estimates are contained within the bounds of Garske et al. who estimates between 51,000 - 380,000 severe infections and 19,000 - 180,000 deaths each year in Africa. We also compare to Shearer et al. who found approximately 256,000 cases on the African continent and 28,000 within Latin America which are at the higher end of our predicted ranges^16^. All the above estimates fall within a similar range despite different scopes and modelling approaches.

In order to produce our burden estimates, we first estimate transmission intensity through a force of infection for each province. These estimates differ to those of Garske et al. in West Africa and the DRC. In order to account for the extended model scope, we revisited the covariates used in the GLM leading to the changes in West African force of infection shown. This can partly be explained by the exclusion of certain covariates, such as longitude, and the inclusion of others, such as NHP species richness. The latter highlights the DRC as an area of high transmission potential, increasing the local estimates of force of infection. The former reduces the force of infection estimates in West Africa. This decrease in West Africa has led to our range of burden lying within the lower range of Garske et al. and whilst the same proportional impact of vaccination is found, the number of deaths averted is also in the lower range of previous estimates. In South America, the force of infections are estimated to be highest in the Amazon region of Brazil, in part due to the high NHP species richness found there. This is consistent with vaccination efforts which have focused on this area leading to relatively low burden despite the high intensity of transmission.

In refining the GLMs of YF occurrence, we expanded the pool of possible covariates. This was partly facilitated by new data becoming available, such as NHP species distributions, and partly motivated by the need to capture more of the inherent variability of YF occurrence, through a temperature suitability index. We find that certain features such as human population size and certain landcover types were consistently featured in the top models of occurrence. Similarly, primate families, Cercopithecidae, Cebidae and Aotidae were all found to be important to YF occurrence. The collection of covariates and model choice lead to high AUCs for each model ranging from 0.935 to 0.949, higher than^17^. One element that is omitted explicitly is vector abundance although we do include occurrence of *Ae. Aegypti* and *Ae. Albopictus*. In recent years there have been a number of excellent efforts to map and predict vector distributions^19,20^. We utilise many of the same model covariates and as such have chosen to omit modelled vector distributions in this analysis. Additionally, YF is transmitted by many vectors whose distributions have yet to be characterised.

There are a number of additional limitations with the range of covariates used. We utilise NHP species presence/ absence data which we aggregate to province level through counts. As such, we do not have information of the population sizes of NHP, only diversity. This could be reassessed as further data becomes available. Additionally, as a component of the covariate selection process, we cluster our covariates based on their correlation with each other. As such, NHPs families who coexist in the same geographic locations are put in the same cluster, eg. Atelidae, leading the model selection process to potentially include a NHP that is present in areas with YF, but not necessarily causing or carrying the virus. As with the other covariates, the selection process implies correlation not causation between the covariate and the occurrence of YF.

Apart from the NHP families, we utilise the same environmental covariates for each continent. Whilst this improves consistency, we use elements such as temperature suitability for *Ae. Aegypti* as a proxy for the vectors of YF and there are different species in each continent which may differ in their own ways to *Ae. Aegypti*. An additional issue is that we aggregate our environmental covariates to province level, reducing resolution and potentially biasing the results. In this study we feel the sparsity of the data, long time window of interest and general uncertainty in other features will eclipse biases introduced by the aggregation mechanism but if this model were to be refined spatially, the bias from aggregation could be readdressed. Finally, whilst we include structural uncertainties from the GLM, we do not included uncertainty in the covariates themselves.

We estimate our models from two main sources of data which we have updated where possible. The occurrence data was expanded to account for additional years and locations from^15,17,21,22^. Yet there are assumptions and uncertainty that are inherent in this data. Firstly, the case definition relies on a vague symptom set which is prone to mis-attribution and may vary by location, this will affect under-reporting. We accommodate variation between countries though country specific reporting factors in the GLMs; however there are number of elements that contribute to surveillance which we essentially aggregate into one component. The most stark difference may be that surveillance in South America uses that the fact that some NHP species experience disease-related mortality as sentinels for YF. In Africa, due to the co-evolution of NHPs and virus, NHPs are not known to be significantly affected in the same way. As such, the surveillance systems are substantially different.

One of the important components to assess inherent under-reporting of YF is serology data. We have significantly expanded the data sources for this aspect of the model, with an addition 36 studies compared to Garske et al. However, all studies are located in Africa, the high vaccination coverage in many of the provinces render conventional tests of seroprevalence useless in terms of assessing background infection. There are further issues that may arise in using seroprevalence. We take a positive serology test to indicate exposure to the disease but also protection, adhering to the conventional assumption that immunity to YF is acquired after infection or vaccination and remains for a lifetime. However, there have been recent studies suggesting this may not be the case in children. Domingo et al. found that immunity against yellow fever waned in children following vaccination^23^. If these results are representative of infant and child vaccination across the regions and time, our estimates of population immunity may need to be readdressed.

Modelling YF is inherently uncertain. We have attempted to quantify this uncertainty through a Bayesian framework and ensemble model predictions. However, there are still elements that we have not captured. Uncertainty in demography and vaccination are not propagated through our model results and yet both will be influential. Demography is captured from UNWPP and scaled according to Landscan. We assume relatively static age structures and, whilst UNWPP goes some way to accounting for population movements, we do not include them explicitly. Population movements not only effect the model directly but will influence the resulting vaccination coverage estimates. In a similar way, vaccination activities are collated from a number of sources with all efforts made to ensure completeness. Yet there activities that may have been omitted or not correctly parameterised which could affect the results of this study. A dominant area of uncertainty is in the symptom spectrum of YF. In our model, we estimate infections and then scale these to arrive at severe infections or cases and, finally, deaths. In this we use the estimates of Johannsson et al. to capture the uncertainty in the proportion of infections considered severe etc.; however, this remains an area of contention for YF as previous estimates of CFR have varied substantially and are significantly larger than other flaviruses such as Dengue^24,25^. The estimates of CFR we use also differ to those in the global burden of disease study leading to substantial differences in burden estimates between the two, although under-reporting is also addressed differently in the GBD^26^.

We have refined an established model but have also inherited some of its’ limitations, one of which is constancy. We assume that dynamics do not change substantially over the observation period in each province. As such, the variation over time is dominated by changes in demography and vaccination. In reality, the epidemiology of YF is likely to change and has seen changes over recent years. Brazil experienced some of the largest outbreaks in its history with YF in 2017 and 2018, this was suggested to have been caused by changing patterns of human behaviour, such as urbanisation and movement, or changes in epidemiology in the sylvatic cycle; however, the full list of causes remains unclear^4,27–30^. Spillover is also inherently stochastic whereas, due to the focus on long-term burden, we assume a constant risk of spillover. As such, the model will not capture outbreak dynamics over a short time window but may highlight areas at most risk of outbreaks. The resulting estimates of burden and vaccine impact are thus the *potential* number of deaths given the conditions in each province and each country given the environmental conditions but may vary year on year due to outbreaks and stochastic spillover events.

## Conclusion

We have refined and extended an established model to update estimates of disease burden for YF. We find consistent results, that 92.2% (95%CrI [88.8%, 95%]) of global burden occurs in Africa and that mass vaccination activities have substantially reduced the number of cases and deaths we see today. We also highlight areas when burden is potentially high, in part due to lower-than-optimal vaccination coverages. As vaccination is the main intervention for YF both as a preventative measure coupled with surveillance, and as an outbreak response intervention, tackling areas of low coverage, including vulnerable age groups within otherwise protected populations, is the optimal route to avert deaths and potential YF outbreaks. However, uncertainty in current data sources, and their interpretation, will limit the effectiveness of planning strategies. Our modelling approach underscores the need to examine background immunity, both due to natural infection and vaccination, in order to address not only the risk of future deaths but to assess how much of YF is actually visible. For an old disease with an effective vaccine, YF still poses new threats, and allowed to run unchecked, will provide a substantial health burden in many tropical areas as well as posing a significant global exportation risk.

## Data Availability

Public repository data:
Vaccination coverage: coverage is available to download from the PoLiCi shiny app: https://shiny.dide.imperial.ac.uk/polici/.
Serology surveys: There are seven published surveys used, available at DOI:10.1016/0147-9571(90)90521-T, DOI:10.1093/trstmh/tru086, DOI: 10.1186/s12889-018-5726-9, DOI: 10.4269/ajtmh.2006.74.1078, PMID: 3501739, PMID: 4004378, PMID: 3731366
Demographic data: Population level data was obtained from UN WPP https://population.un.org/wpp/, this was disaggregated using Landscan 2014 data https://landscan.ornl.gov/landscan-data-availability.
Environmental data: This was obtained from LP DAAC: https://lpdaac.usgs.gov/ and worldclim http://www.worldclim.org/
Yellow fever outbreaks: These were compiled from the WHO weekly epidemiologic record and disease outbreak news https://www.who.int/wer/en/ and https://www.who.int/csr/don/en/.
Data elsewhere:
The data from the WHO YF surveillance database and from recent serological surveys from WHO member states in Africa underlying the results presented in the study are available from World Health Organization (contact: William Perea, or Laurence Cibrelus, or Jennifer Horton). Data from the occurrence of YF in Brazil were obtained from the Brazilian MoH (contact: Daniel Garkauskas Ramos).

https://shiny.dide.imperial.ac.uk/polici/

## Contributors

KAMG, ATPH, NMF and TG conceptualised the study. KAMG wrote the original draft. TG, ATPH, KAMG, LC and DGR compiled datasets. KAMG and ATPH did the analysis. All authors contributed to reviewing and editing of the manuscript and have approved the final version.

