## Supplementary Information for "The global burden of yellow fever"

### The global burden of yellow fever: supplementary material

04 October, 2020

#### 1 Data

We combine multiple data sets within a Bayesian framework to account for areas with sparse data and under-reporting. The model is estimated at admin level one, or province, level to match the available occurrence data.

###### 1.0.1 Global yellow fever occurrence

A database of yellow fever occurrence was collated. This was compiled in two parts. Occurrence in Africa was compiled originally in Garske et al. and has been subsequently maintained and updated [8, 10, 15]. Occurrence of yellow fever in South America was collated by Hamlet and colleagues [12]. Reports of yellow fever in humans were assembled for both continents from sources including the weekly epidemiological record [30], disease outbreak news [29], WHO yellow fever surveillance database (YFSD), Brazilian ministry of health (MoH) and Pan-American Health Organisation (PAHO). Provinces reporting at least one case of YF since 1984 are shown in figure 1 in the main text.

###### 1.0.2 Vaccination coverage and demography

We use the methodology of Garske et al. [8] with updated data sets and additional data for South America in order to estimate vaccination coverage across the regions. The results of this methodology are visualised in finer detail, admin level 2, in the POLICI shiny application where the coverage is available to download from 1940 to 2050 [11]. The coverage is informed by historic data on mass-vaccination activities, reactive campaigns, recent preventive mass vaccination campaigns and routine infant vaccination [4, 22, 31]. Further data was provided by the MoH for Brazil.

Demographic data was obtained from the United Nations World population prospects (UNWPP) which provides country level population sizes [26]. These were dis-aggregated to province level using Landscan data on population distributions [3, 18]. Age distributions were assumed to be the same across all provinces and population distributions were assumed not to substantially vary over the observation period. Landscan provides population size estimates at 1/120 degree resolution. Combining this with UNWPP, we arrive at the total number of individuals in each age group and province over time.

###### 1.0.3 Environmental data

The generalised linear model of yellow fever occurrence was developed to account for dependence on environmental conditions, habitat suitability and occurrence of the non-human primates (NHPs). A full list of potential covariates are available in the supplementary material. Covariates include measures of enhanced vegetation index (EVI), altitude, temperature, precipitation and land cover types as well as non-human primate (NHP) species richness eg. Atelidae, occurrence of *Ae. Aegypti* and *Ae. Albopictus*, and *Ae. Aegypti* temperature suitability [5, 23, 32, 16].

Table S1: Composition of the 20 best fitting generalised linear models of YF reports. Surveillance quality is also included in all models.

| Model variant | A | B | C | D | E | F | G | H | I | J | K | L | M | BIC |
|---|---|---|---|---|---|---|---|---|---|---|---|---|---|---|
| 1 | 1 | 1 | 1 | 1 | 1 | 0 | 0 | 1 | 1 | 0 | 0 | 1 | 0 | 870 |
| 2 | 1 | 1 | 1 | 1 | 1 | 1 | 1 | 1 | 1 | 1 | 1 | 1 | 1 | 872 |
| 3 | 1 | 1 | 1 | 1 | 1 | 0 | 0 | 1 | 1 | 0 | 0 | 1 | 1 | 872 |
| 4 | 1 | 1 | 1 | 1 | 1 | 1 | 1 | 1 | 1 | 1 | 1 | 0 | 0 | 872 |
| 5 | 1 | 1 | 1 | 1 | 1 | 1 | 1 | 1 | 1 | 1 | 1 | 0 | 1 | 873 |
| 6 | 1 | 1 | 1 | 1 | 1 | 1 | 1 | 1 | 1 | 0 | 0 | 1 | 0 | 873 |
| 7 | 1 | 1 | 1 | 1 | 1 | 1 | 1 | 1 | 1 | 1 | 1 | 1 | 0 | 873 |
| 8 | 1 | 1 | 1 | 1 | 1 | 1 | 1 | 1 | 1 | 0 | 0 | 1 | 1 | 873 |
| 9 | 1 | 1 | 1 | 1 | 1 | 1 | 0 | 1 | 1 | 0 | 0 | 1 | 0 | 873 |
| 10 | 1 | 1 | 1 | 1 | 1 | 1 | 1 | 1 | 1 | 1 | 0 | 1 | 1 | 874 |
| 11 | 1 | 1 | 1 | 1 | 1 | 1 | 1 | 1 | 1 | 0 | 1 | 1 | 0 | 874 |
| 12 | 1 | 1 | 1 | 1 | 1 | 1 | 1 | 1 | 1 | 0 | 1 | 1 | 1 | 874 |
| 13 | 1 | 1 | 1 | 1 | 1 | 1 | 1 | 1 | 1 | 1 | 0 | 1 | 0 | 875 |
| 14 | 1 | 1 | 1 | 1 | 1 | 1 | 1 | 0 | 1 | 0 | 0 | 1 | 0 | 875 |
| 15 | 1 | 1 | 1 | 1 | 1 | 0 | 0 | 0 | 1 | 0 | 0 | 1 | 0 | 875 |
| 16 | 1 | 1 | 1 | 1 | 1 | 0 | 0 | 1 | 1 | 1 | 0 | 1 | 0 | 875 |
| 17 | 1 | 1 | 1 | 1 | 1 | 1 | 1 | 1 | 1 | 0 | 1 | 0 | 0 | 875 |
| 18 | 1 | 1 | 1 | 1 | 1 | 0 | 0 | 1 | 1 | 0 | 1 | 1 | 0 | 875 |
| 19 | 1 | 1 | 1 | 1 | 1 | 1 | 0 | 1 | 1 | 0 | 0 | 1 | 1 | 875 |
| 20 | 1 | 1 | 1 | 1 | 1 | 1 | 1 | 0 | 1 | 1 | 1 | 0 | 0 | 876 |

Covariate data sets were available as gridded datasets of various spatial resolution. These were aggregated to province level, the same scale as the occurrence data. Temperature, altitude and precipitation data was obtained from Worldclim version 2.0 [5]. These were aggregated by calculating the mean, max or min over the area of the province. Land cover was obtained from MODIS [7, 6]. This was aggregated by examining the proportion of each province coverage by a landcover type. NHP species distributions were obtained from the IUCN red list [13]. These were aggregated to province level by counting the species richness in each area. Species occurrence was considered if the species range covered more than 10% of the admin unit. Occurrence of *Ae. Aegypti* and Albopictus was obtained from the supplementary information of Kraemer et al. [16].

Prior to fitting, all variables were scaled to unit variance.

###### 1.0.4 Serological surveys

We use serological surveys to assess transmission intensity in specific regions. Unfortunately, these are only available in the African endemic region. We include all surveys included in Gaythorpe et al. [10] as well as a newly published survey undertaken in Kenya [1, 2, 17, 20, 24, 25, 28]. In the majority of surveys, individuals known to have been vaccinated are omitted; however, in south Cameroon this information is unavailable and so we estimate an additional vaccination factor. In the study of Chepkorir et al. we include vaccinated proportions as stated in their evaluation after omitting those with unknown status.

#### 2 Environmental covariates

A full list of covaraites is provided in table S3.

Table S2: Covariate definitions

| Covariate | Meaning |
|---|---|
| A | NHP Cercopithecidae species richness |
| B | NHP Cebidae species richness |
| C | log(Population) |
| D | Temperature suitability (mean) |
| E | Grasslands land cover |
| F | Savanna land cover |
| G | Evergreen broadleaf forests |
| H | Aedes Aegypti Occurrence |
| I | NHP Aotidae species richness |
| J | Woody savanna land cover |
| K | Temperature range |
| L | Maximum middle infrared reflectance (MIR) |
| M | Altitude |

Table S3: Full list of environmental covariates included.

| | | | | |
|---|---|---|---|---|
| logpop | MIR.min | LC9 | albopictus | family_HOMINIDAE |
| temp_min | MIR.max | LC10 | altitude | family_INDRIIDAE |
| temp_mean | LC1 | LC11 | family_AOTIDAE | family_LEMURIDAE |
| precip_max | LC2 | LC12 | family_ATELIDAE | family_LEPILEMURIDAE |
| precip_min | LC3 | LC13 | family_CALLITRICHIDAE | family_LORISIDAE |
| precip_mean | LC4 | LC14 | family_CEBIDAE | family_PITHECIIDAE |
| EVI.mean | LC5 | LC15 | family_CERCOPITHECIDAE | EVI.range |
| EVI.min | LC6 | LC16 | family_CHEIROGALEIDAE | temp_range |
| EVI.max | LC7 | LC17 | family_DAUBENTONIIDAE | temp_suit_mean |
| MIR.mean | LC8 | aegypti | family_GALAGIDAE | temp_suit_min |

Table S4: Temperature suitability index parameter values.

| | $a_c$ | $a_{T_0}$ | $a_{T_m}$ | $\rho_c$ | $\rho_{T_0}$ | $\rho_{T_m}$ | $\mu_c$ | $\mu_{T_0}$ | $\mu_{T_m}$ |
|---|---|---|---|---|---|---|---|---|---|
| Value | 2.72e-4 | 2.24 | 40.13 | -0.75 | 12.71 | 38.05 | 1.36e-4 | 17.33 | 42.20 |

#### 2.1 Non-human primates

Non-human primate data was acquired from the IUCN redlist [13]. This provided presence and absence data at species level. In order to produce maps of primate species richness, we perform a count of all species belonging to a family in each province.

#### 2.2 Temperature suitability

Temperature suitability was calculated as Gaythorpe et al. [9]. The form of the temperature suitability index is given by:

$$z(T) = \frac{a(T)^2 \exp(-\mu(T)\rho(T))}{\mu(T)}$$

where the bite rate, extrinsic incubation period and mosquito mortality, given by $a, \rho$ and $\mu$ respectively, are affected by temperature $T$ in the following ways:

$$\begin{aligned} a(T) &= a_c T(T - a_{T_0})(a_{Tm} - T)^{0.5} \\ \rho(T) &= 1/(\rho_c T(T - \rho_{T_0})(\rho_{Tm} - T)^{0.5}) \\ \mu(T) &= 1/(-\mu_c(T - \mu_{T_0})(\mu_{T_m} - T)) \end{aligned}$$

following [21]. The subscripts $c, 0$ and $m$ represent the positive rate constant, min temperature and max temperature for each thermal response model. These were estimated in Gaythorpe et al. [9] within a Bayesian framework and we retain the point estimates shown in table S4.

#### 2.3 Models

We extend the model of Garske et al. to account for YF burden in South America as well as Africa [8]. We update currently included data and include further data where necessary to expand the scope of the modelling.

##### 2.3.1 Seroprevalence

We assume a constant force of infection for each province over the observation period. This is the same as Garske et al. and was found to be a better reflection of available data by Gaythorpe et al. [10] than that another, dynamic, model variant. We assume homogeneous mixing and account for vaccination using the following form for $s(\lambda, u)$, the expected seroprevalence in age group $u$ given force of infection $\lambda$:

$$s(\lambda, u) = 1 - \left(1 - \frac{\sum_{a \in u}(1 - exp(-\lambda a)p_a)}{\sum_{a \in u}(p_a)}\right)\left(1 - \frac{\sum_{a \in u} v_a p_a}{\sum_{a \in u} p_a}\right),$$

where $a$ indexes the annual age groups, $p_a$ is the population age distribution and $v_a$ is the vaccination coverage in age group a. The binomial log likelihood is then given by the following:

$$\log L_{sero} = \sum_u \log \binom{N_u}{K_u} s(\lambda, u)^{K_u} (1 - s(\lambda, u))^{N_u}),$$

where $N_u$ is the number of samples in age group u and $K_u$ is the number of positive samples in age group u.

###### 2.3.2 Generalised linear model of the presence/ absence of yellow fever reports

A generalised linear model was fitted to the dataset of yellow fever occurrence from 1984 to 2019 at province level. The data is assumed to be binomially distributed and a complementary log-log link function is used such that model predictions in province i, $q_i$, are given by

$$q = 1 - exp(-e^{X\beta}),$$

where $X$ denotes the matrix of covariates and $\beta$ indicates the parameter vector to be fitted. The log-likelihood is given by

$$\log L_{glm} = \sum_i (y_i \log(q_i) + (1 - y_i)\log(1 - q_i)),$$

where $y_i$ denotes the presence/ absence in province i.

The occurrence of yellow fever depends on a number of environmental factors as well as the abundance and distribution of the vector and NHP hosts. We consider many of these variables as potential covariates in the model. As with Garske et al., the number of covariates to consider is large, and has been extended for the current work by the inclusion of NHPs and temperature suitability. As such we perform a selection process, detailed in full in the supplementary material and summarised below:

1. We remove covariates that are not significantly associated with the data. For each covariate, we fit a univariate GLM to the data using the base R function, `glm`. We remove covariates with p-value $< 0.1$; in this case, all covariates were significant.
2. Highly correlated covariates are clustered such that the pairwise correlation in each cluster exceeds 0.75. This produces 38 clusters.
3. We choose one covariate from each cluster to be further examined. Here, the covariate with the maximum absolute correlation with the data is chosen.
4. The function `stepAIC` from the `MASS` package is used to further whittle the list of covariates down [27]. We choose the multiplier of the number of degrees of freedom such that the test criterion is BIC: the Bayesian information criterion instead of AIC: the Akaike information criterion.
5. The final step is to use the R package, `bestglm` to produce the best 20 models according to BIC [19]. This uses the complete enumeration algorithm.

All models then included a measure of surveillance quality. For the 21 countries within the yellow fever surveillance database, specific data on reporting per capita was available. For countries not covered by the yellow fever surveillance database, and thus without an independent estimate of surveillance, individual country factors were fitted. However, countries not considered at risk were grouped together in order to have one country factor. This is in order to avoid infinite parameter estimates in areas which are known not to have yellow fever reports.

###### 2.3.3 Transmission intensity

The transmission intensity estimates arising from the serology allow us to calculate the number of infections over the observation period in the areas where surveys were conducted. We link this to the probability of yellow fever report through a Poisson reporting process with a probability of the detection. This is calculated by comparing the GLM to predictions of the seroprevalence models in the following way:

$$q_i = 1 - (1 - \rho_i)^{n_{inf,i}},$$

where $\rho_c$ is the per-country probability of detection, $q_i$ is the probability of a report in province $i$, provided by the GLM, and $n_{inf,i}$ is the number of infections in province $i$, provided by the seroprevalence model. This means that the probability of detection can be linked to the GLM covariates by:

$$n_{inf,i} \log(1 - \rho_c) = \exp(X\beta),$$

and in terms of the country factors, GLM covariates $\beta_c$, and $b$, the baseline surveillance quality by:

$$\log(-\log(1 - \rho_c)) = \beta_c + b.$$

Once the probability of detection has been estimated for each province with a serological study, we take the mean over each and use the resulting probability to extrapolate transmission intensity in areas where there are currently no seroprevalence studies.

#### 2.4 Estimation

The best fitting models, according to BIC, were estimated within a Bayesian framework described in Gaythorpe et al. including code used. The estimation is divided into two phases. The GLMs are estimated using adaptive Markov Chain Monte Carlo (MCMC) sampling whereas the seroprevalence models are estimated within the product space framework of Gaythorpe et al. [10] with the probability of the force of infection model set to 1; as such, the estimation becomes an adaptive MCMC with log-transformed parameters.

Prior distributions were chosen in many cases to match those of Garske et al. [8]. Country factors retain the Gaussian prior distribution with mean 0 and standard deviation 2 except for the countries considered low risk, whose country factor had truncated normal prior with mean 0 and standard deviation 30 and limits $[0, \infty)$. The same prior was used for the GLM coefficient for `aggregate_family`, the aggregated NHP species richness and the converse for the GLM coefficients for temperature range and altitude which were assumed to be negative. All other GLM coefficient priors were normal with mean 0 and standard deviation 30. The force of infections for each seroprevalence study had exponential priors with rate parameter 0.001 and the vaccine efficacy had truncated normal prior with mean 0.975 and standard deviation 0.05, according to Jean et al. [14]; this was truncated to [0,1].

#### 2.5 Ensemble predictions

We propagate uncertainty from both the parameter estimation and model structure. This is done through sampling proportionally from the posterior distributions of all 20 of the best-fitting models to produce 500 force of infection and thus burden predictions. We sample proportional to the AUC of each model fit.

##### 2.5.1 Estimation diagnostics

###### 2.5.1.1 Generalised linear models

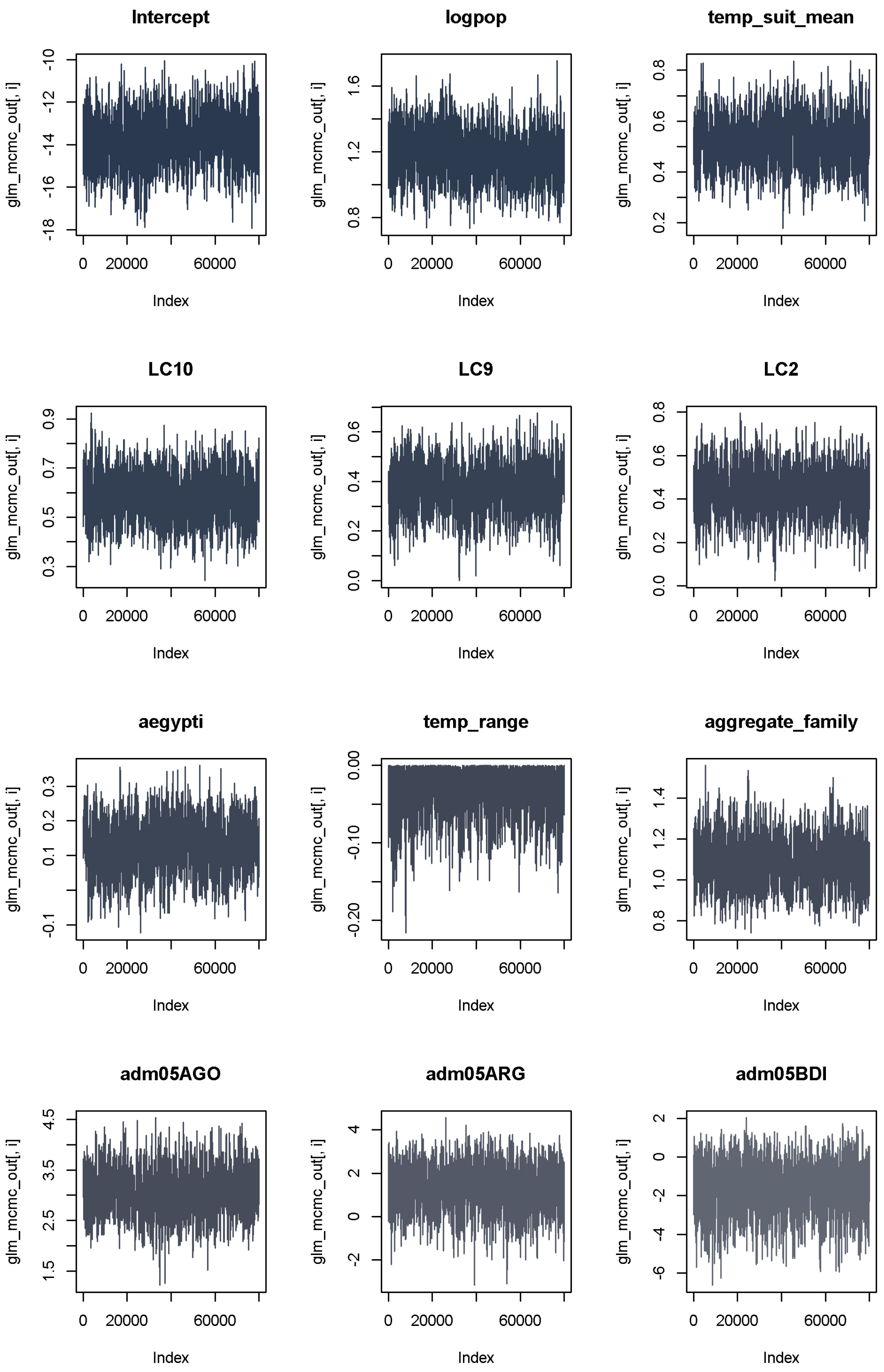


Figure S1: Trace plots from estimation of model variant 17 as an example of convergence.

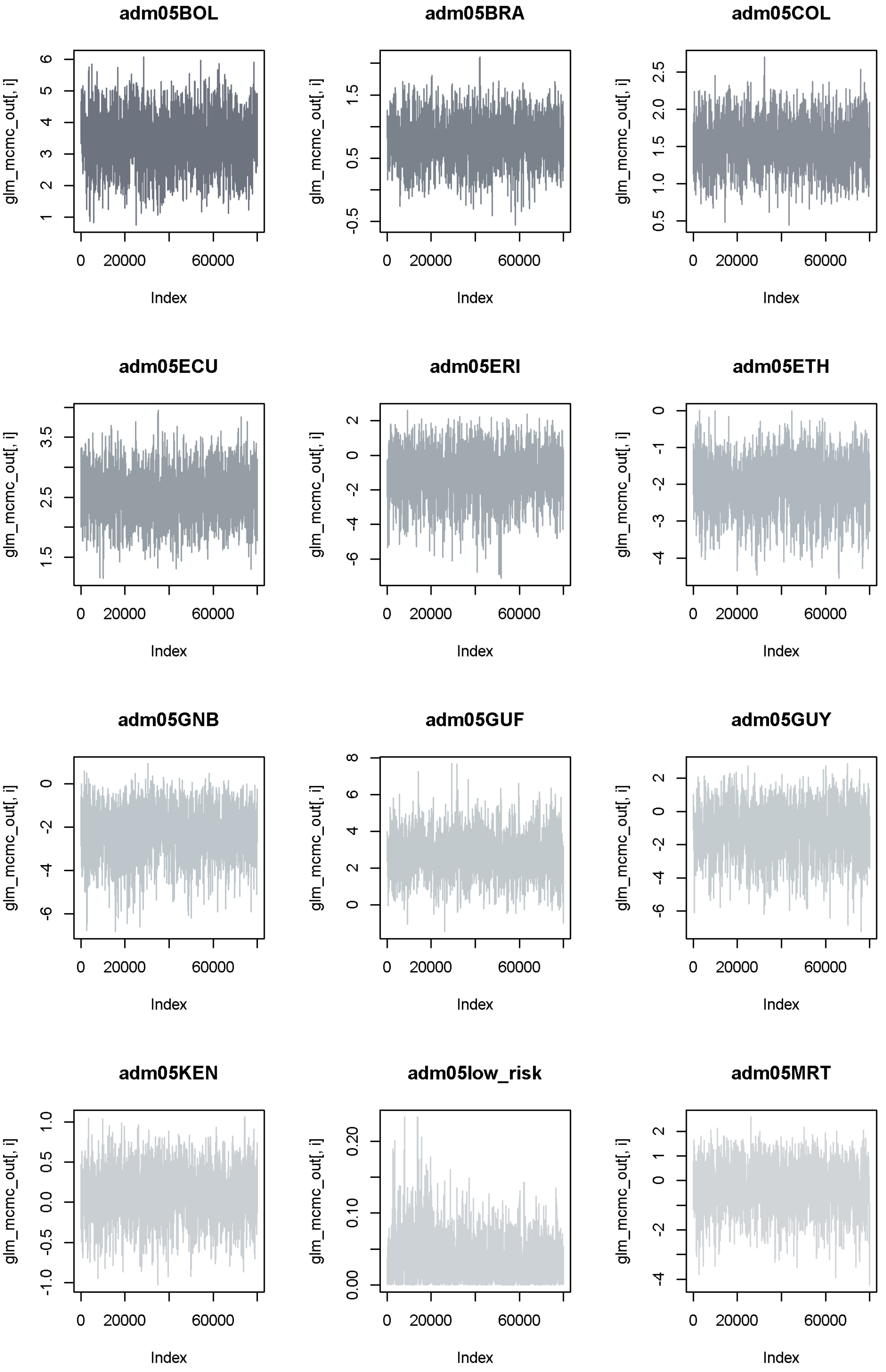


Figure S2: Trace plots from estimation of model variant 17 as an example of convergence.

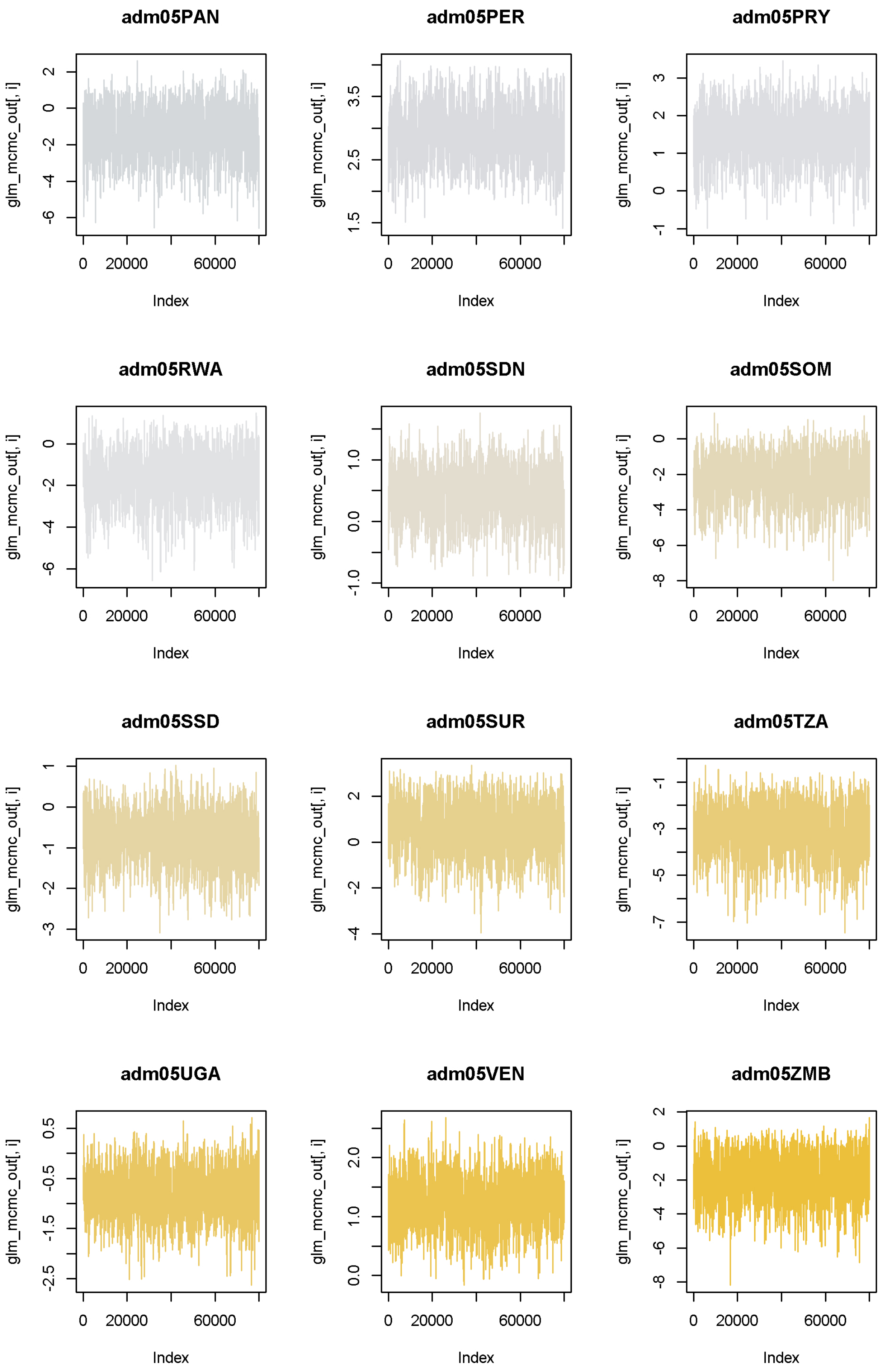


Figure S3: Trace plots from estimation of model variant 17 as an example of convergence.

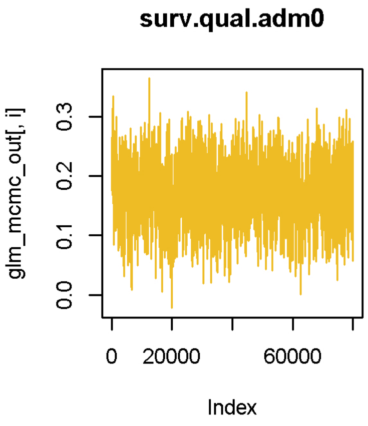


Figure S4: Trace plots from estimation of model variant 17 as an example of convergence.

###### 2.5.1.2 Seroprevalence models

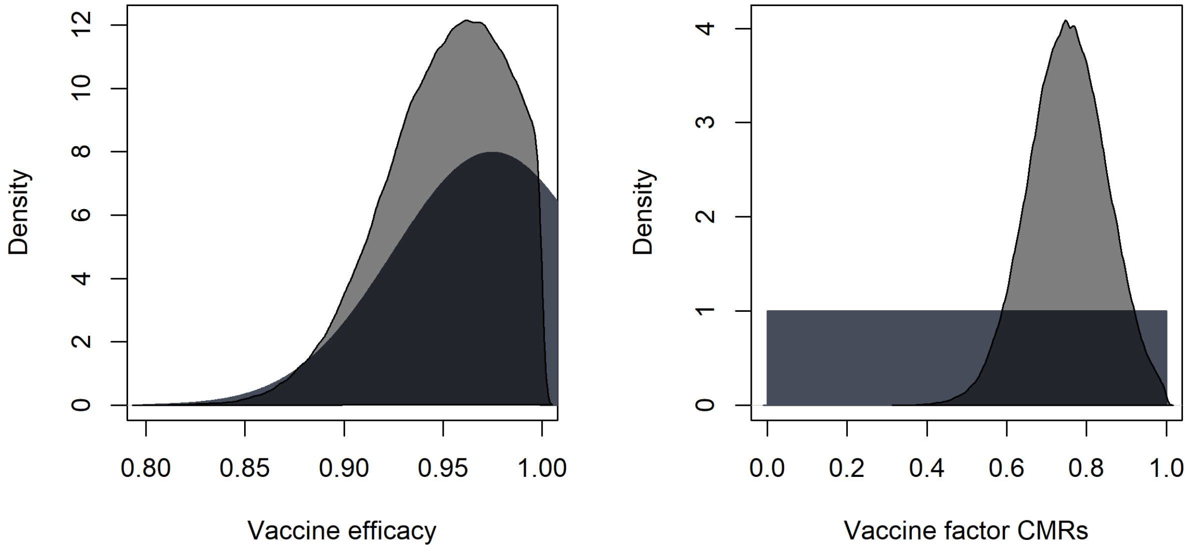


Figure S5: Prior and posterior distributions for vaccine efficacy and vaccine factor for CMRs.

### 3 Uncertainty in transmission intensity projections

We calculate the coefficient of variation for all provinces with respect the 100 samples of force of infection for each of the 20 best-fitting glms.

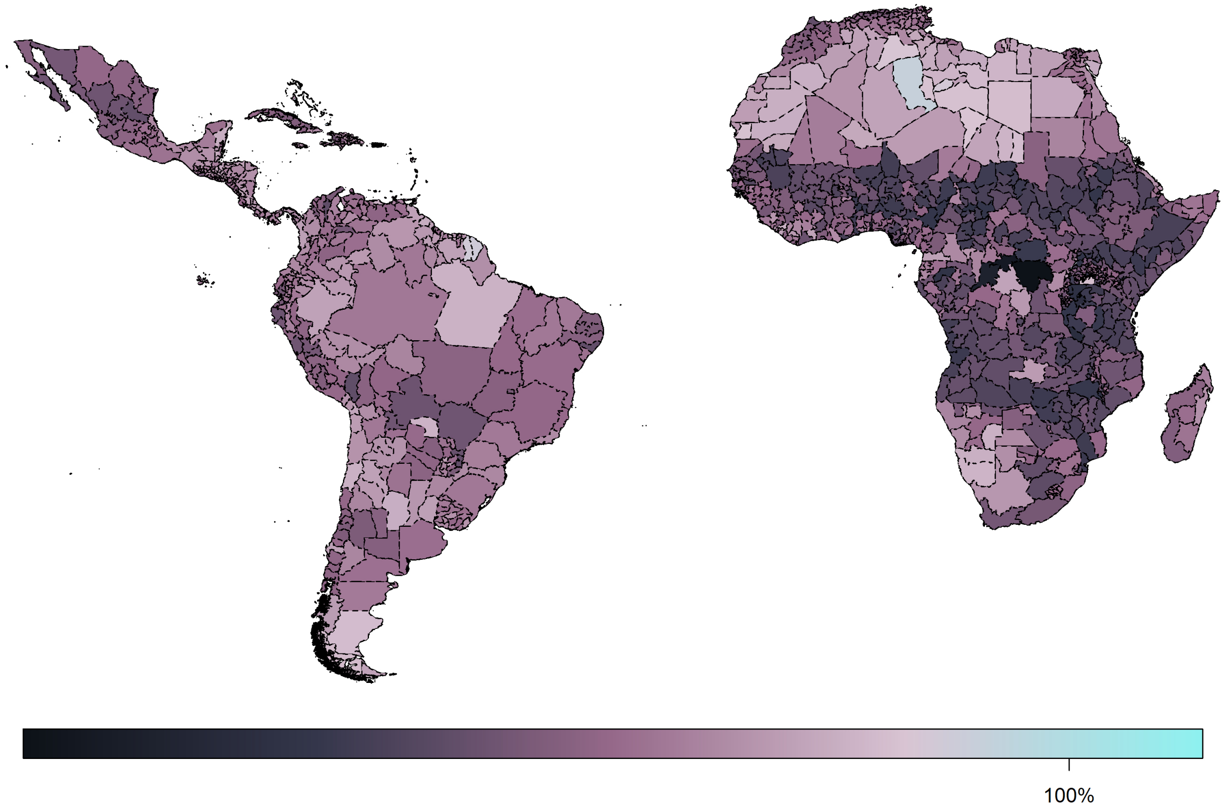


Figure S6: Coefficient of variation in the force of infection estimates between 100 samples of each of the 20 best models.

#### 4 Impact of mass vaccination in 2013

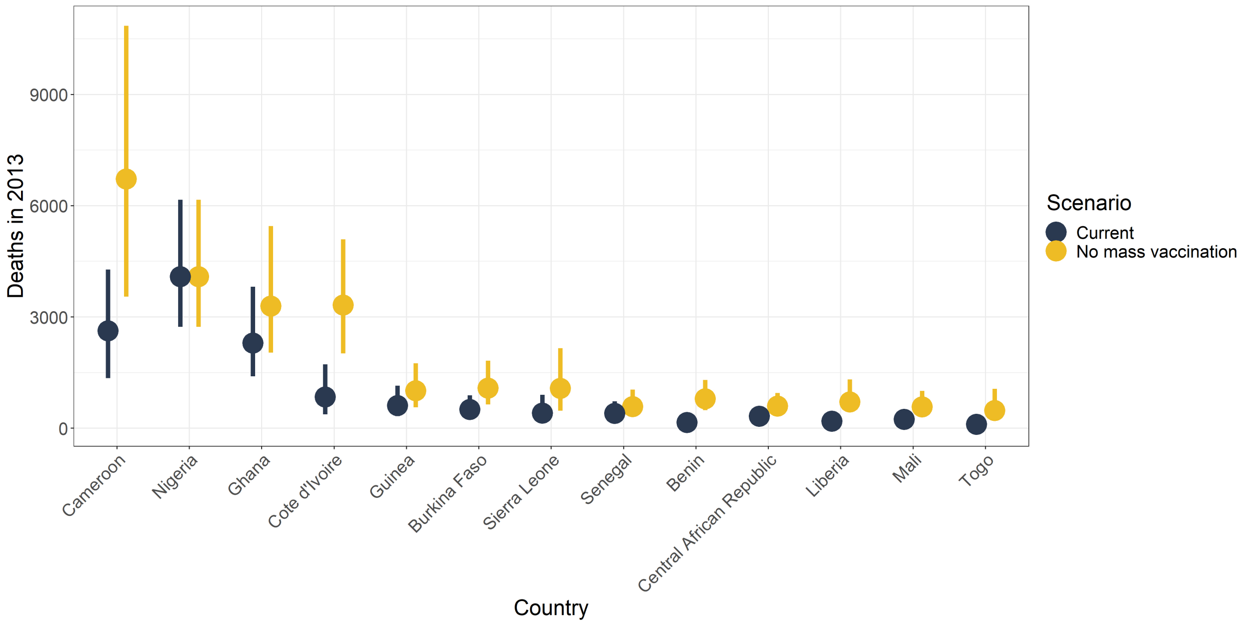


Figure S7: Median posterior predicted deaths averted for 2013 by country. Yellow represents the number of deaths without mass vaccination campaigns since 2006 and black represents deaths with current vaccination coverage levels. The mid line denotes median and the box range shows the 95% credible interval.
